# Reducing Information and Selection Bias in EHR-Linked Biobanks via Genetics-Informed Multiple Imputation and Sample Weighting

**DOI:** 10.1101/2024.10.28.24316286

**Authors:** Maxwell Salvatore, Ritoban Kundu, Jiacong Du, Christopher R Friese, Alison M Mondul, David Hanauer, Haidong Lu, Celeste Leigh Pearce, Bhramar Mukherjee

## Abstract

Electronic health records (EHRs) are valuable for public health and clinical research but are prone to many sources of bias, including missing data and non-probability selection. Missing data in EHRs is complex due to potential non-recording, fragmentation, or clinically informative absences. This study explores whether polygenic risk score (PRS)-informed multiple imputation for missing traits, combined with sample weighting, can mitigate missing data and selection biases in estimating disease-exposure associations. Simulations were conducted for missing completely at random (MCAR), missing at random (MAR), and missing not at random (MNAR) conditions under different sampling mechanisms. PRS-informed multiple imputation showed generally lower bias, particularly when combined with sample weighting. For example, in biased samples of 10,000 with exposure and outcome MAR data, PRS-informed imputation had lower percent bias (3.8%) and better coverage rate (0.883) compared to PRS-uninformed (4.5%; 0.877) and complete case analyses (10.3%; 0.784) in covariate-adjusted, weighted, multiple imputation scenarios. In a case study using Michigan Genomics Initiative (n=50,026) data, PRS-informed imputation aligned more closely with a sample-weighted All of Us-derived benchmark than analyses ignoring missing data and selection bias. Researchers should consider leveraging genetic data and sample weighting to address biases from missing data and non-probability sampling in biobanks.

---

Electronic health records (EHRs) represent a rich, longitudinal resource that researchers increasingly use to address questions of public health and clinical significance. EHR-linked biobanks, which often contain genetic information linked to other data sources (e.g., administrative and insurance claims, neighborhood-level characteristics, and complementary survey data), are growing in both the number of participants (n) and the breadth of measured variables (p). However, EHR data has not been collected for research purposes, so researchers must carefully consider potential biases (i.e., systematic errors). Potential sources of bias include missing data^1–3^ (including clinically informative visiting processes^2,4,5^), selection bias,^6–8^ misclassification,^7,9,10^ confounding,^11,12^ immortal time bias,^13,14^ and clinical practice and data collection and processing heterogeneity across EHRs.^15,16^ Although the advent of large-scale secondary data (colloquially, “Big Data”^17^) effectively minimizes the threat of random error, systematic sources of bias are ever-present adversaries, unphased by ever-increasing sample sizes. In fact, large sample sizes amplify these biases relative to the very small variance, frequently making inference erroneous, a phenomenon commonly characterized as the Big Data Paradox.^18^

Missing data is ubiquitous in epidemiology^19–23^ and almost universally encountered in health research.^24,25^ Complete case analyses, which ignore observations with missing data for variables of interest (e.g., exposures, outcomes, or important covariates), are the most commonly employed approach in randomized clinical trials^25^ and observational studies^24^ in the presence of missing data. However, complete case analyses can lead to biased parameter estimation depending on the missing data mechanism, or the reason why the data are missing.^1,19,26^

Missing data mechanisms broadly fall into three classes: missing completely at random (MCAR), missing at random (MAR), and missing not at random (MNAR).^27^ Naïve complete-case analyses are expected to produce unbiased results when data are MCAR. However, there are several reasons that make the assumption that missing data are MCAR in EHR-linked biobanks less reasonable,^6,30–32^ including non-random patient-provider interactions,^33–35^ clinically informative observation processes,^2,36^ and EHR fragmentation. For these reasons, MAR and MNAR assumptions are more plausible.^37^ Among the existing missing data methods that improve precision and reduce bias (e.g., inverse probability weighting^38–40^ and full-information maximum likelihood^41–44^), multiple imputation is a commonly used and frequently recommended approach for handling missing data in EHR-linked biobanks.^3,37,45–47^ It is important to note that MNAR data cannot be empirically distinguished from MAR data while the MCAR assumption can be tested.^48,49^

A hallmark of major biobanks is availability of genetic data on a large fraction of participants and an active genetics research community producing polygenic risk scores (PRS) for a variety of traits.^50,51^ It is an interesting question whether PRS observed on a large sample can improve imputation of the traits they are constructed for. The actual traits may be missing for a large number of participants, and PRS can serve as a weak proxy.^52,53^

Adding to the missing data challenge is the fact that EHR-linked biobanks often do not represent their source (or target) population, introducing potential selection bias.^54^ Recruitment mechanisms like recruiting patients awaiting surgery (as in the Michigan Genomics Initiative (MGI)^55^) and oversampling groups historically underrepresented in biomedical research (as in the NIH All of Us Research Program^56^) as well as participant-driven factors like healthy volunteer bias (as in the UK Biobank^57,58^) explain differences between the study cohorts and their underlying source and target populations.^57,59^ Weighting-based methods like inverse probability weighting and poststratification weighting are often employed to reduce selection bias in parameter estimation when individual or summary data from an external non-probability sample are available. Recent papers have shown that weighted analyses reduce (but do not remove) bias due to selection in EHR-linked biobanks.^58,60,61^

In this study we considered, to the best of our knowledge, an unexplored question: can PRS-informed multiple imputation reduce bias due to missing exposure data in association estimation? We investigated (a) whether PRS-informed multiple imputation meaningfully reduces bias due to missing data in probability samples and (b) the joint impact of PRS-informed multiple imputation and sample weighting on exposure-outcome association estimation in biased samples (Figure 1). We calculated unweighted and weighted complete case- and multiple imputation-based estimates of the body mass index (BMI) coefficient for glucose in realistic simulations, followed by a case study stratified by non-Hispanic White (n=42,999) and non-Hispanic Black (n=2,297) status in MGI. First, our simulation studies explored the joint impacts of multiple imputation with and without exposure and outcome PRS for missing data in MCAR, MAR, and MNAR settings and weighting in random and biased samples. We considered sampling weights in biased samples. Our case study applied these methods to MGI data to estimate the BMI coefficient for glucose using the same missing data methods and stratum-specific selection weights (as described previously^60^) to demonstrate differences in association estimates under different analytical strategies in real-world data relative to National Health Interview Survey-weighted All of Us-based benchmark.

**Figure 1.**
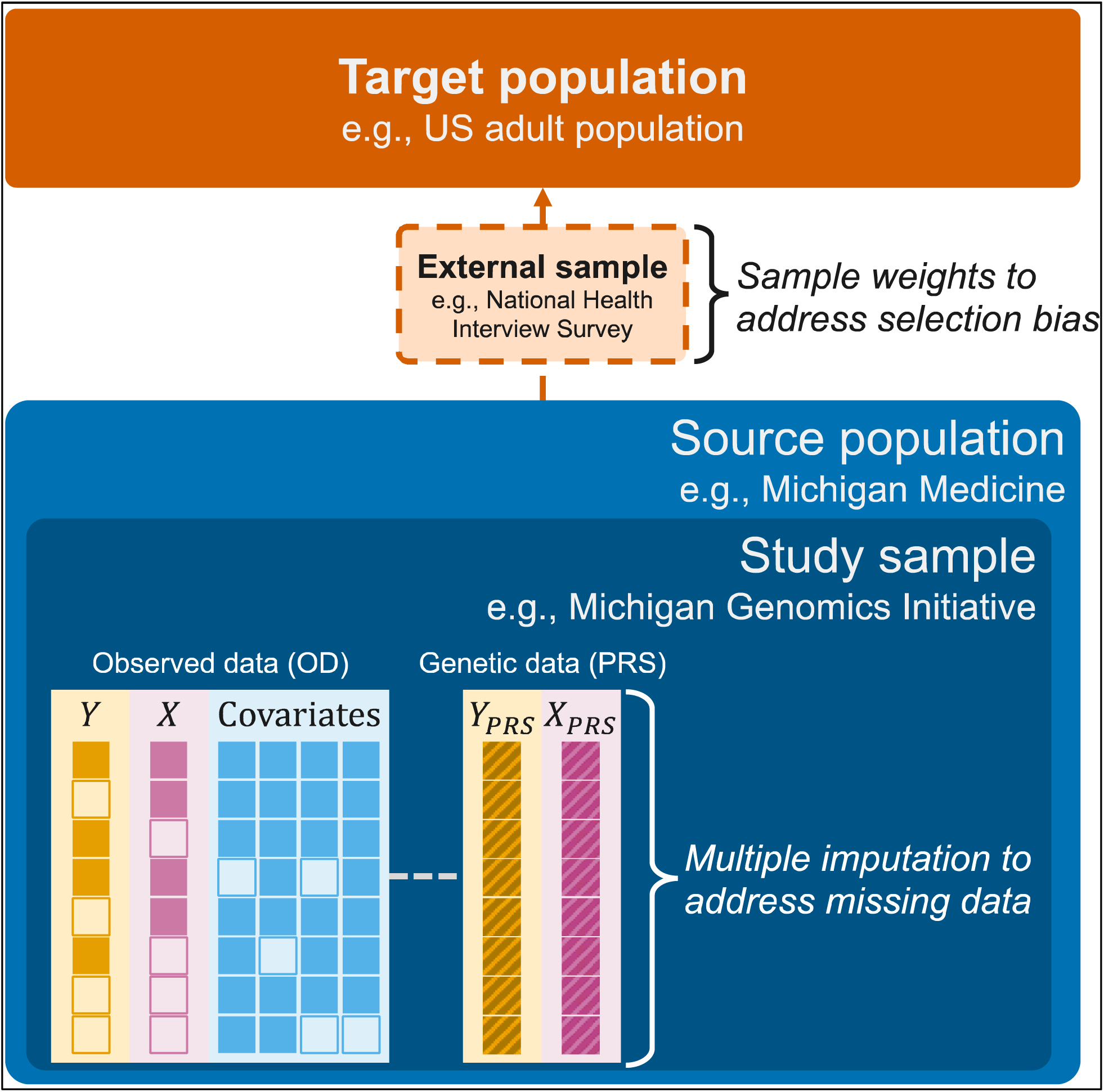
Schematic representation depicting multiple imputation and weighted analyses to jointly address missing data and selection bias. *Y* represents the outcome (e.g., glucose), *X* represents the exposure (e.g., body mass index) and covariates could include age, sex, race/ethnicity, and smoking status. The empty boxes represent missing data. *Y_PRS_* and *X_PRS_* are the polygenic risk scores (PRS) corresponding to the outcome and exposure, respectively.

## METHODS

### Simulation Design

#### Generating outcome, exposure, covariates, and polygenic risk scores jointly

We simulated 1,000 replicates of a pseudo-population with size 100,000 (Figure 2). To achieve this, we first generated an 8-dimensional multivariate normal distribution, *X*∼*N*_8_(*µ*, *t*), mimicking the joint distribution of age, sex, non-Hispanic White (NHW), smoking status (ever/never), BMI, glucose, BMI PRS, and glucose PRS, assuming mean standardized variables (µ= 0) and t as observed in MGI (see Eq.1 below).

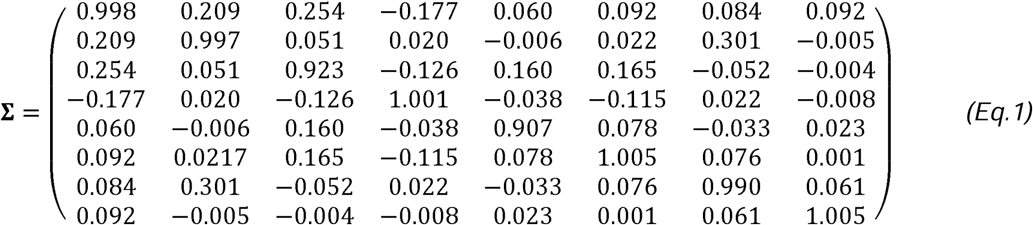

**Figure 2.**
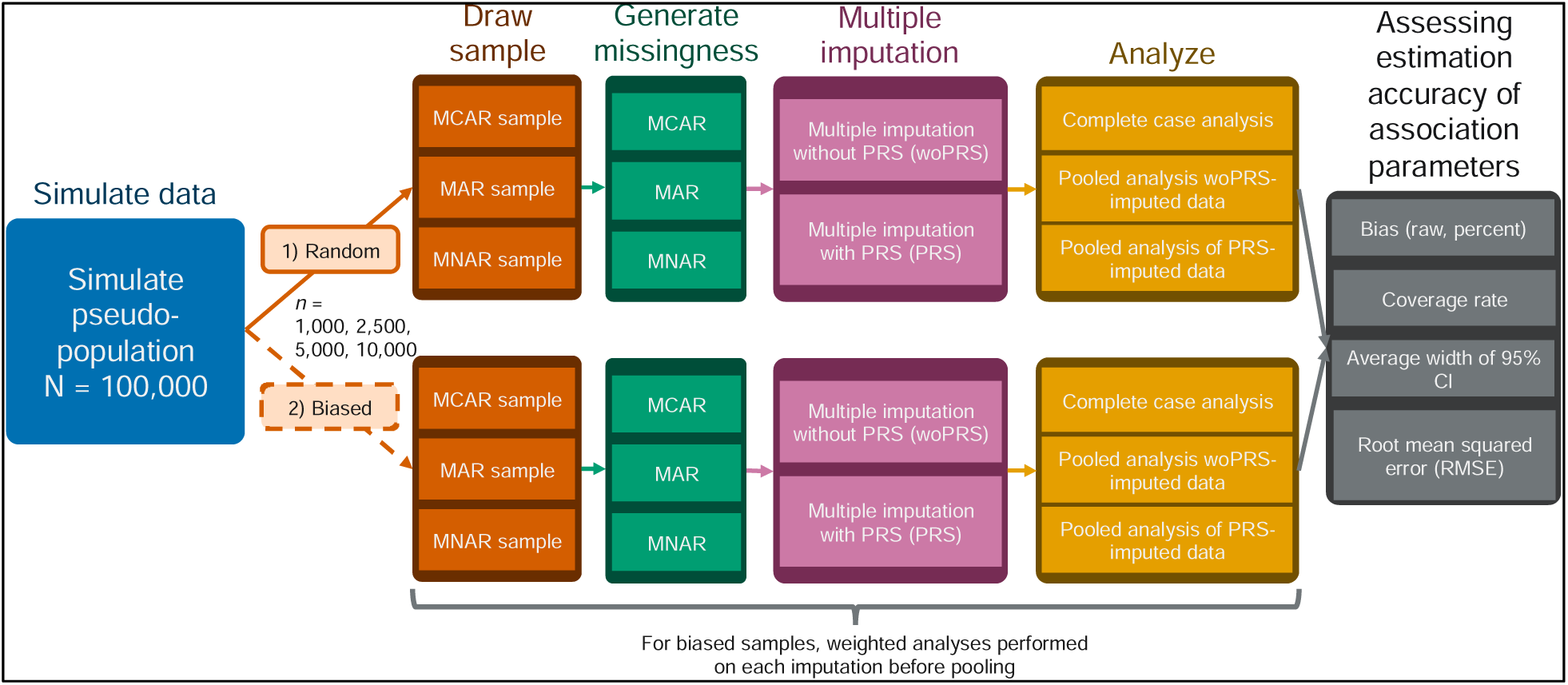
Schematic representation of random and biased sampling simulation analyses. Abbreviations: CI, confidence interval; MAR, missing at random; MCAR, missing completely at random; MNAR, missing not at random; PRS, polygenic risk score; RMSE, root mean square error; woPRS, without polygenic risk score

Binary variables were recoded (sex, NHW, smoking status) from the generated continuous variables such that they preserved their correlation with age in observed MGI data.

#### Sample selection

For each pseudo-population, we performed sampling under two scenarios: random and biased/covariate-informed. Covariate-informed sampling probabilities depended on observed age, glucose (the outcome), and BMI (the exposure) (e.g., *logi (P(s= 1|age, glucose, BMJ)) = y_O_ + y_age_age + y_glucose_glucose + y_BMI_ BMJ*, where s is an indicator variable for selection into the sample; *y_age_, y_glucose_, y_BMI_* = 1). For each scenario, the intercept, *y_O_*, was selected to draw a sample of approximately 1,000, 2,500, 5,000, and 10,000 (y_O_ = -6.92, -5.83, -4.94, -3.93, respectively) from the pseudo-population where individual *i* had selection probability *P(s_i_ = 1|age_i_, glucose_i_, BMJ_i_))* dependent on the exposure (BMI) and the outcome (glucose) as well as the covariate (age).

#### Missingness generation

We simulated (a) exposure only (i.e., BMI) and (b) exposure and outcome (i.e., BMI and glucose) missingness under MCAR, MAR, and MNAR mechanisms for each selected sample size and mechanism. Approximately 25% missingness was generated for each variable. Under MCAR, the probability of missingness (e.g*., P(R_BMI_* = 1) where *R_BMI_* is an indicator for whether BMI is missing) was 25% for all observations. Under MAR, exposure missingness depended on the outcome (glucose) and covariates (age, sex, race/ethnicity, smoking status) while outcome (glucose) missingness depended only on covariates. Under MNAR, exposure and outcome missingness was dependent on the whole set of exposure, outcome, and covariates. In all settings, all non-intercept regression coefficients were set equal to 1 and only the intercept was tuned to attain the desired sample size. Supplementary Table 1 shows the intercept coefficient values by missingness mechanism to (approximately) achieve the desired sample sizes.

#### Analytic Methods

For each scenario, we performed unweighted and weighted analyses (for simple random sampling, these are equivalent). The weights were proportional to the inverse of the *known* covariate-informed sampling probabilities for each individual *i* (*w_i_ o< P(s_i_ = 1|age_i_, glucose_i_, BMJ_i_)_-_*_1_), and weighted analyses were carried out using the survey R package (version 4.4-2).^62^ In addition to complete case analysis, we performed multiple imputation to address missing data. For each sample, multiple imputation methods using age, sex, NHW, smoking status, BMI, glucose (without PRS; woPRS-imputed) and additionally exposure (BMI) and outcome (glucose) PRS (PRS-imputed) were carried out using the R package mice (version 3.16.0; m = 5 imputations).^63,64^ Beta coefficients across imputations were pooled using Rubin’s rule, with confidence intervals calculated from pooled standard errors based on within and between imputation variances.^65,66^ For multiply imputed analyses of biased samples, weighted analyses were conducted on each imputed dataset before pooling.

Our target quantity was the true coefficient of BMI in a linear regression model for glucose (β*_BMI_*) adjusted for age, sex, NHW race/ethnicity, and smoking status (ever/never) (Eq. 2).

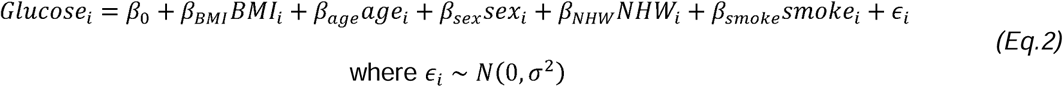

For each replicate, the true β*_BMI_* was obtained from the pseudo-population of size 100,000 and the sample estimates were obtained in the selected samples of sizes 1,000, 2,500, 5,000, and 10,000. In each sample, we conducted unweighted and weighted complete case, woPRS-imputed, and PRS-imputed analyses, extracting the coefficient estimate of BMI for glucose (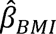*_BMI_*). We evaluated association estimation properties using percent bias, coverage rate, average 95% confidence interval width, and root mean square error (RMSE), averaged over the 1,000 replicates.

### Case study: Michigan Genomics Initiative

#### Description of the study cohort

MGI is an EHR-linked biobank that began in 2012, initially recruiting adult patients through pre-/peri-operative appointments requiring anesthesia from the University of Michigan Health System. As of September 2023, ∼100,000 consented participants have provided access to their EHR and a biospecimen for genotyping, with a recent follow-up effort collecting complementary survey data.^67^ This paper included 42,999 (25,520 complete cases) non-Hispanic White and 2,297 (1,240 complete cases) non-Hispanic Black participants aged 40 or older without a diabetes diagnosis and with demographic, health measurement, laboratory, and polygenic risk score data. MGI protocols were reviewed and approved by the University of Michigan Medical School Institutional Review Board (IRB ID HUM00099605 and HUM00155849).

#### Outcome, exposure, covariates, and polygenic risk score

The outcome and exposure of interest were glucose (mg/dL; logical observation identifiers names and codes (LOINC) code: 2345-7) and BMI (kg/m^2^), respectively. The longitudinal data in the EHR was reduced to the participant’s median value after removing extreme values (values outside 1.5x the interquartile range) for the corresponding variable. Age was considered the participant’s age at the time of data pull (March 23, 2022). Sex (indicator for female) and race/ethnicity were obtained from EHR data. Multiple measurements of self-reported smoking status were recorded and recoded into a binary ever/never indicator variable.

Ma and colleagues previously calculated several PRS for 27 exposures in MGI participants.^51^ In this paper, we selected the Lassosum PRS for BMI and the deterministic Bayesian sparse linear mixed model PRS for glucose because they had the highest R^2^ value for their respective traits in the published paper.^51^ These PRSs relied on publicly available GWAS summary statistics of UK Biobank data (Neale lab^68,69^). Both PRSs were predictive in MGI, with BMI PRS being much stronger (Pearson correlation between BMI and BMI PRS: 0.30; glucose and glucose PRS: 0.09; Supplementary Figure 1).

#### Estimated regression coefficient corresponding to BMI with glucose as the outcome

The target estimand of interest was the regression coefficient corresponding to BMI with glucose as the outcome. We conducted analyses among individuals 40 and older without a diabetes diagnosis in non-Hispanic White and non-Hispanic Black strata as well as in the full (i.e., unstratified) cohort. Unlike in the simulations, the selection weights in MGI were *not known*. Salvatore and colleagues estimated inverse probability selection weights to make MGI more representative of the US adult population using National Health Interview Survey data.^60^ Using the same methods to calculate stratum-specific weights, we conducted weighted versions of each regression analysis. Using the non-Hispanic White and non-Hispanic Black samples with missing data (n=42,999 and 2,297, respectively), we performed multiple imputation with and without PRS (adjusting for age, sex, and smoking status). We also conducted a PRS-informed multiple imputation analysis where observations were restricted to only those with observed PRS (PRS-imputed (subset): n=25,520 and 1,240 for non-Hispanic Whites and non-Hispanic Blacks, respectively). We reported the estimated beta coefficients and 95% confidence intervals.

### Software

Analyses were conducted using R version 4.3.3. The code used to conduct analyses in this paper is available at https://github.com/maxsal/exprs_imputation.

## RESULTS

### Simulation study

In **random sampling with exposure-only missingness**, when BMI data was MCAR (Figure 3), all analyses successfully maintained the nominal 95% coverage rates and exhibited no bias in the estimated BMI coefficients for glucose. However, when the data was MAR, complete case analysis showed a decline in coverage rates as sample size increased, and consistently exhibited bias (e.g., 8.86% for n=1,000; 7.80% for n=10,000). woPRS-imputed analyses, in contrast, provided more stable coverage rates (e.g., 0.931 for n=1,000; 0.924 for n=10,000) and reduced bias (e.g., 2.89% for n=1,000; 0.10% for n=10,000) as sample sizes grew. PRS-imputed analyses outperformed both, fully retaining the nominal coverage rate across all sample sizes and achieving the least bias (e.g., 1.5% for n=1,000; 0.04% for n=10,000). Under MNAR conditions, none of the analyses could maintain the nominal coverage rate, and all exhibited substantial bias exceeding 30%. However, PRS-imputed analyses performed slightly better than woPRS-imputed analyses, achieving marginally higher coverage rates (e.g., for n=1,000: 0.637 for PRS-imputed; 0.561 for woPRS-imputed) and lower bias (e.g., for n=1,000: 31.95% for PRS-imputed; 36.12% for woPRS-imputed).

**Figure 3.**
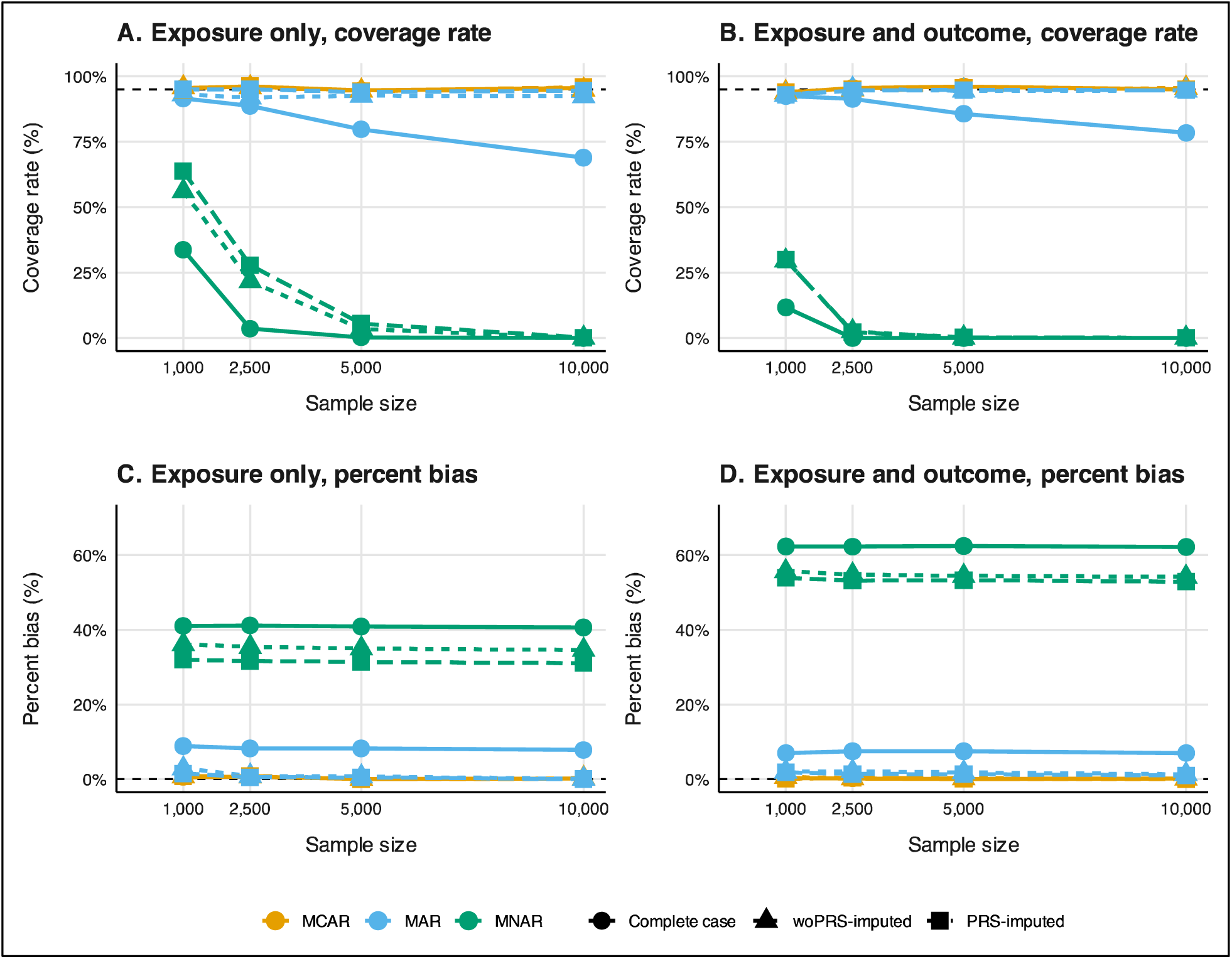
Coverage rate (panels A and B) and percent bias (panels C and D) diagnostics for exposure only (panels A and C) and exposure and outcome missingness (panels B and D) BMI coefficient for glucose by missing data mechanism and method and sample size under random sampling in a 1,000-iteration simulation. Analyses were adjusted for age, sex, non-Hispanic White, and smoking status (ever/never). Corresponding coverage rate, percent bias, average confidence interval width, and root mean squared error diagnostics are reported in Supplementary Table 2 and Supplementary Table 3. Abbreviations: MAR, missing at random; MCAR, missing completely at random; MNAR, missing not at random; PRS-imputed, polygenic risk score-informed multiple imputation; woPRS-imputed, multiple imputation without exposure and outcome PRS.

In **biased sampling scenarios where only exposure data were missing**,MCAR conditions led to significant bias and a failure to retain nominal coverage across all unweighted analyses, with the bias worsening as sample sizes increased (e.g., >29%; Figure 4). When the missingness was MAR or MNAR, unweighted complete case analyses exhibited less bias than multiple imputation methods, likely due to the amplification of biases by multiple imputation when it does not account for sampling weights. However, when analyses were weighted, multiple imputation approaches, particularly PRS-imputed, showed substantial improvements in coverage rates and bias reduction, consistently outperforming complete case analyses. For instance, in a 10,000-observation sample with MAR missingness, the coverage rate was 0.784 for complete case analysis, 0.877 for woPRS-imputed, and 0.883 for PRS-imputed. When data was MNAR, coverage decreased, and bias increased across all methods, but PRS-imputed analyses performed slightly better, with reduced bias as sample size increased.

**Figure 4.**
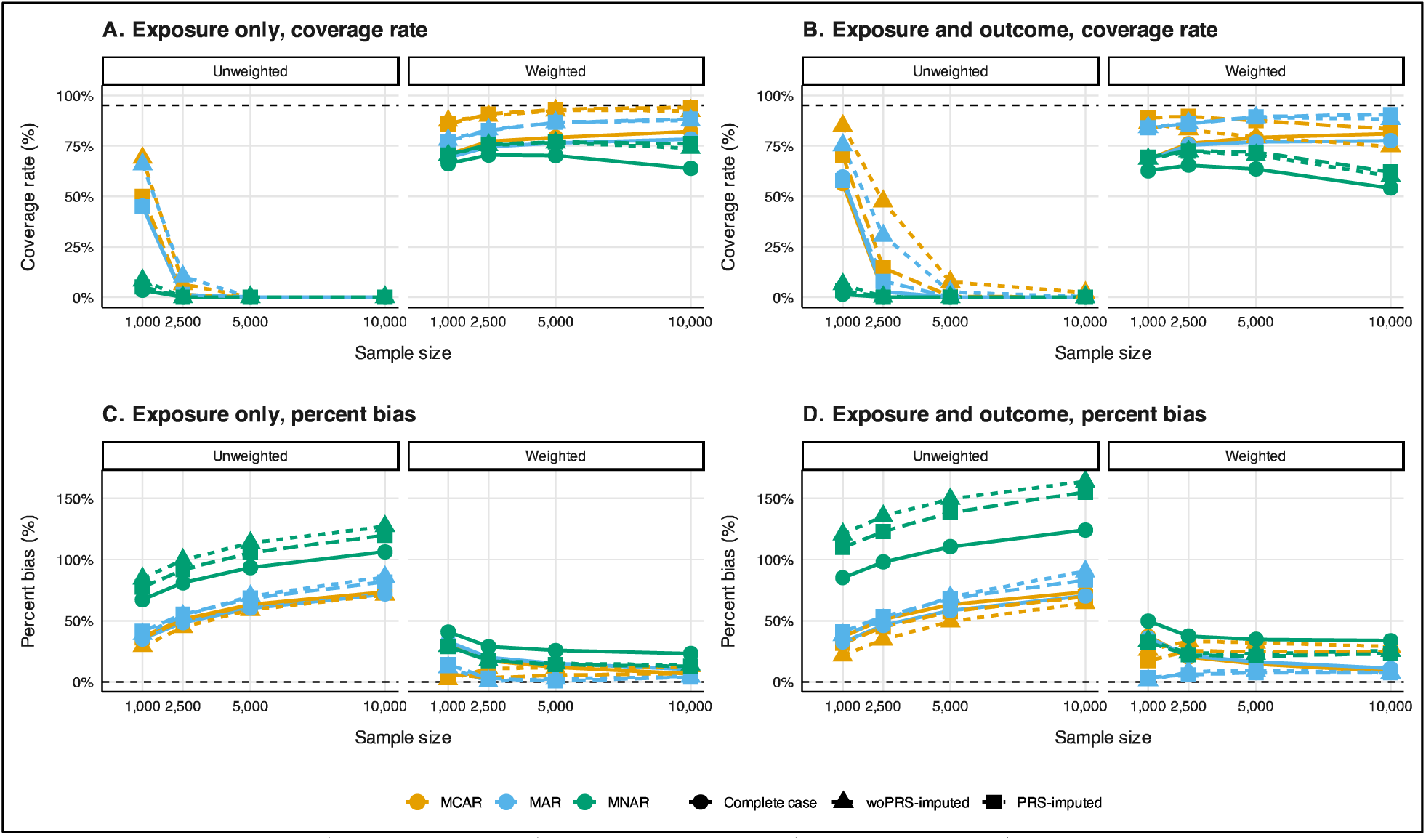
Coverage rate (panels A and B) and percent bias (panels C and D) diagnostics for unweighted (left) and weighted (right) BMI coefficient for glucose estimation by missing data mechanism and method and sample size under biased sampling and exposure only (panels A and C) and exposure and outcome missingness (panels B and D) in a 1,000-iteration simulation. For biased sampling simulations, unweighted and weighted diagnostics are reported in Supplementary Tables 4, 5, 6, and 7, respectively. Analyses were adjusted for age, sex, non-Hispanic White, and smoking status (ever/never).

In **random sampling scenarios where both exposure and outcome data were missing,** MCAR conditions allowed all analyses to maintain the nominal coverage rates and remain unbiased (Figure 3). For example, the coverage rate for complete case analysis was 0.938 for n=1,000 and remained above 0.950 for larger sample sizes. However, under MAR conditions, coverage rates decreased, and bias remained stable with larger sample sizes (e.g., coverage rate dropped from 0.925 for n=1,000 to 0.784 for n=10,000) in complete case analyses. woPRS-imputed and PRS-imputed analyses effectively returned to nominal coverage rates in MAR data (e.g., 0.946 and 0.947, respectively, for n=10,000) and exhibited little to no bias (e.g., 1.32% for woPRS-imputed; 0.97% for PRS-imputed). Under MNAR conditions, no analysis method could maintain nominal coverage, and all showed significant bias. Nonetheless, PRS-imputed analyses slightly outperformed others, achieving better coverage rates (e.g., 0.299 for PRS-imputed versus 0.000 for complete case in n=1,000) and lower bias (e.g., 53.89% for PRS-imputed versus 62.27% for complete case in n=1,000). Plots describing average 95% CI width and RMSE are shown in Supplementary Figure 2.

In **biased sampling with both exposure and outcome missingness**, unweighted PRS-imputed analyses failed to recover nominal coverage and exhibited significant bias across all missingness mechanisms, with MCAR data showing the least bias (Figure 4). When sampling weights were applied, PRS-imputed multiple imputation analyses performed better than complete case analyses, with improved coverage rates, particularly in MAR data. For example, PRS-imputed coverage rates for MAR data exceeded those observed for MCAR as sample sizes increased (e.g., 0.840 for n=1,000 to 0.906 for n=10,000). Despite these improvements, MNAR data analyses remained problematic across all methods, with PRS-imputed analyses showing slightly better performance but still exhibiting considerable bias and suboptimal coverage (e.g., percent bias of 32.40% for n=1,000 and 23.19% for n=10,000; plots depicting average 95% CI width and RMSE are shown in Supplementary Figures 3 and 4).

### Analysis in the Michigan Genomics Initiative (MGI)

#### Descriptive characteristics of the study population

We analyzed a cohort of 50,026 MGI participants aged 40 or older without diabetes, of which 54.5% were female and 86.0% non-Hispanic White. The mean age was 62.9 years (SD: 12.5), BMI was 29.1 (6.0), and glucose was 99.0 mg/dL (14.1) (Supplementary Table 8). Due to suspected racial/ethnic heterogeneity,^70,71^ we stratified the analysis into non-Hispanic White and non-Hispanic Black groups.

Among the 42,999 non-Hispanic White participants, 53.8% were female, with a mean age of 63.5 years (12.5), BMI of 29.1 (6.0), and glucose of 99.5 mg/dL (14.2) (Table 1). Participants with missing data were generally younger (mean age 62.6 vs. 64.1 years), more likely to be female (54.6% vs. 53.3%), less likely to have smoked (46.5% vs. 50.7%), and had slightly lower glucose levels (99.47 vs. 99.55 mg/dL), with no differences in BMI PRS (p=0.6) or glucose PRS (p=0.4) compared to those with complete data.

**Table 1.**
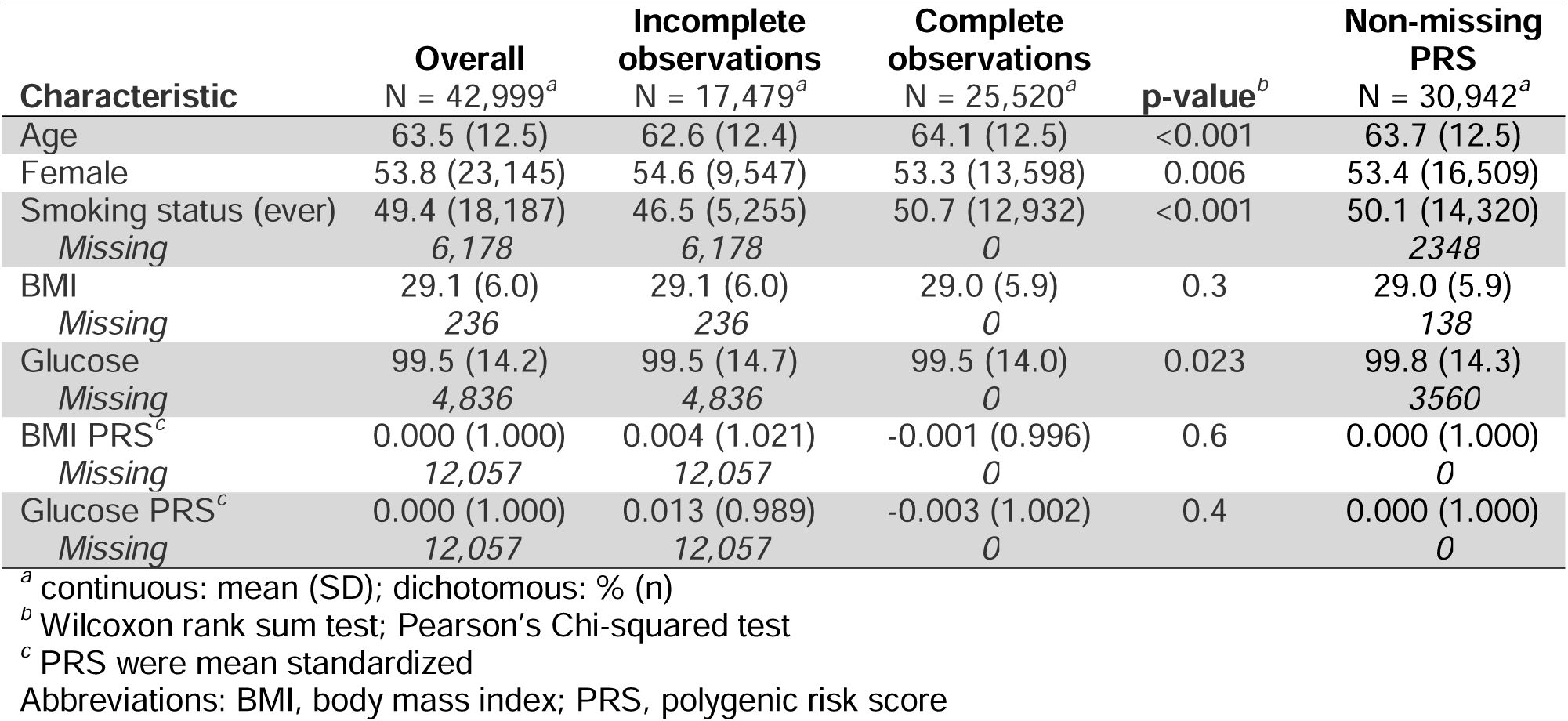
Comparison of demographic, health measurements, and polygenic risk score values overall and among non-Hispanic Whites 40 or older without diabetes, with and without any missing values in the Michigan Genomics Initiative.

| Characteristic | Overall<br>N = 42,999<sup>a</sup> | Incomplete<br>observations<br>N = 17,479<sup>a</sup> | Complete<br>observations<br>N = 25,520<sup>a</sup> | p-value<sup>b</sup> | Non-missing<br>PRS<br>N = 30,942<sup>a</sup> |
| --- | --- | --- | --- | --- | --- |
| Age | 63.5 (12.5) | 62.6 (12.4) | 64.1 (12.5) | <0.001 | 63.7 (12.5) |
| Female | 53.8 (23,145) | 54.6 (9,547) | 53.3 (13,598) | 0.006 | 53.4 (16,509) |
| Smoking status (ever) | 49.4 (18,187) | 46.5 (5,255) | 50.7 (12,932) | <0.001 | 50.1 (14,320) |
| Missing | 6,178 | 6,178 | 0 |  | 2348 |
| BMI | 29.1 (6.0) | 29.1 (6.0) | 29.0 (5.9) | 0.3 | 29.0 (5.9) |
| Missing | 236 | 236 | 0 |  | 138 |
| Glucose | 99.5 (14.2) | 99.5 (14.7) | 99.5 (14.0) | 0.023 | 99.8 (14.3) |
| Missing | 4,836 | 4,836 | 0 |  | 3560 |
| BMI PRS<sup>c</sup> | 0.000 (1.000) | 0.004 (1.021) | -0.001 (0.996) | 0.6 | 0.000 (1.000) |
| Missing | 12,057 | 12,057 | 0 |  | 0 |
| Glucose PRS<sup>c</sup> | 0.000 (1.000) | 0.013 (0.989) | -0.003 (1.002) | 0.4 | 0.000 (1.000) |
| Missing | 12,057 | 12,057 | 0 |  | 0 |

<sup>a</sup> continuous: mean (SD); dichotomous: % (n)
<sup>b</sup> Wilcoxon rank sum test; Pearson's Chi-squared test
<sup>c</sup> PRS were mean standardized
Abbreviations: BMI, body mass index; PRS, polygenic risk score

Among the 2,297 non-Hispanic Black participants, 63.3% were female, with a mean age of 57.8 years (11.4), BMI of 30.8 (6.4), and glucose of 95.1 mg/dL (12.5) (Table 2). Those with missing data were less likely to have smoked (39.2% vs. 44.7%) and had lower glucose levels (94.0 vs. 95.8 mg/dL), with no differences in BMI PRS (p=0.8) or glucose PRS (p=0.061) compared to complete cases.

**Table 2.** Comparison of demographic, health measurements, and polygenic risk score values overall and among non-Hispanic Blacks 40 or older without diabetes, with and without any missing values in the Michigan Genomics Initiative.

| Characteristic | Overall<br>N = 2,297<sup>a</sup> | Incomplete<br>observations<br>N = 1,057<sup>a</sup> | Complete<br>observations<br>N = 1,240<sup>a</sup> | p-value<sup>b</sup> | Non-missing<br>PRS<br>N = 1,437<sup>a</sup> |
| --- | --- | --- | --- | --- | --- |
| Age | 57.8 (11.4) | 57.3 (11.2) | 58.2 (11.6) | 0.069 | 57.7 (11.6) |
| Female | 63.3 (1,454) | 63.2 (668) | 63.4 (786) | >0.9 | 63.3 (910) |
| Smoking status (ever) | 42.6 (847) | 39.2 (293) | 44.7 (554) | 0.017 | 44.3 (597) |
| Missing | 310 | 310 | 0 |  | 88 |
| BMI | 30.8 (6.4) | 30.6 (6.3) | 31.0 (6.5) | 0.2 | 31.0 (6.5) |
| Missing | 26 | 26 | 0 |  | 8 |
| Glucose | 95.1 (12.5) | 94.0 (12.6) | 95.8 (12.4) | <0.001 | 95.8 (12.4) |
| Missing | 160 | 160 | 0 |  | 116 |
| BMI PRS<sup>c</sup> | 0.000 (1.000) | -0.006 (0.977) | 0.001 (1.004) | 0.8 | 0.000 (1.000) |
| Missing | 860 | 860 | 0 |  | 0 |
| Glucose PRS<sup>c</sup> | 0.000 (1.000) | 0.158 (1.064) | -0.025 (0.988) | 0.061 | 0.000 (1.000) |
| Missing | 860 | 860 | 0 |  | 0 |

<sup>a</sup> continuous: mean (SD); dichotomous: % (n)
<sup>b</sup> Wilcoxon rank sum test; Pearson's Chi-squared test
<sup>c</sup> PRS were mean standardized
Abbreviations: BMI, body mass index; PRS, polygenic risk score

Subsets of 30,492 non-Hispanic Whites and 1,437 non-Hispanic Blacks had complete PRS data. Smoking status and glucose showed moderate missingness (14% and 11% in the non-Hispanic White sample and 13% and 7% in the non-Hispanic Black, respectively), while BMI was rarely missing in both groups (0.5% in non-Hispanic Whites and 1.1% in non-Hispanic Blacks).

#### Estimation of the coefficient for BMI with glucose as the outcome

Among non-Hispanic White individuals aged 40 years or older without diabetes in the MGI cohort, the unweighted, covariate-adjusted, complete case coefficient estimate was 0.288 (0.264, 0.312) (Figure 5), which differed from the benchmark range of 0.376 to 0.423, derived from NHIS-weighted All of Us data. Using sampling weights, the estimate improved to 0.324 (0.283, 0.365), aligning more closely with the benchmark. Multiple imputation alone also improved the estimates, with woPRS-imputed and PRS-imputed estimates rising to 0.300 (0.277, 0.323) and 0.302 (0.280, 0.324), respectively. When these imputation methods were combined with sample weighting, they approached the benchmark even more closely, with final estimates of 0.331 (0.292, 0.371) for woPRS-imputed and 0.338 (0.280, 0.324) for PRS-imputed analyses. *Overall, sample weighting and multiple imputation consistently brought estimates closer to the benchmark*. Similar trends were seen in the corresponding unadjusted analyses in Figure 5A.

**Figure 5.**
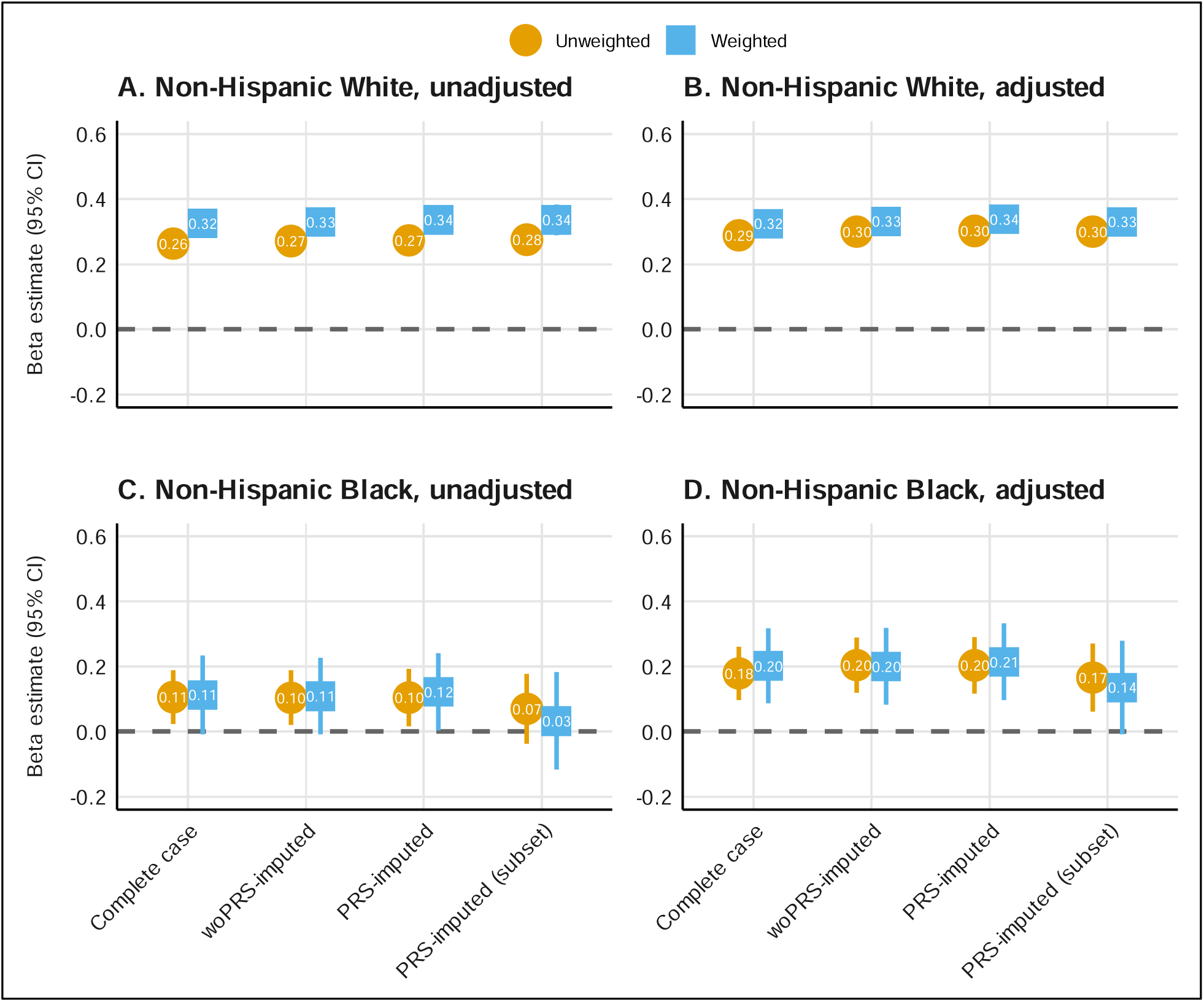
Estimation of the coefficient for BMI with glucose as the outcome by missing data method and weighting approach among non-Hispanic Whites (n=42,999; panels A and B) and non-Hispanic Blacks (n=2,297; panels C and D) in all MGI adults age 40 or older without diabetes. The PRS-imputed subset sample (n=30,942 for non-Hispanic Whites; n=1,437 for non-Hispanic Blacks) was restricted to individuals with non-missing genotype data before multiple imputation. Analyses were adjusted for age, sex, and smoking status (ever/never). Gray shaded regions represent corresponding 95% confidence interval from National Health Interview Survey-weighted All of Us data where weights are calculated separately for non-Hispanic Whites and non-Hispanic Blacks to make All of Us data for each of these groups more representative of their corresponding US population (target population). Results for the full, unstratified cohort are shown in Supplementary Figure 5. Abbreviations: PRS, polygenic risk score.

In non-Hispanic Black individuals aged 40 years or older without diabetes, the unweighted complete case estimate was 0.178 (0.097, 0.261) (Figure 5), also differing from the benchmark range of 0.196 to 0.297. However, applying sampling weights brought the estimate within the benchmark range at 0.202 (0.086, 0.317). Multiple imputation alone saw nominal increases in estimates, with woPRS-imputed and PRS-imputed estimates at 0.204 (0.119, 0.288) and 0.203 (0.116, 0.290), respectively. The weighted analyses produced similar results, with estimates of 0.200 (0.082, 0.318) for woPRS-imputed and 0.214 (0.096, 0.332) for PRS-imputed analyses. Unlike the non-Hispanic White group, weighting had a smaller impact because the estimates were already within the benchmark range.

In the unstratified results for the entire MGI cohort aged 40 years or older without diabetes (Supplementary Figure 5), which was predominantly non-Hispanic White (86%), the findings mirrored those of the non-Hispanic White stratum. For example, the weighted PRS-imputed estimate (0.312 (0.274, 0.349)) was closer to the benchmark range of 0.346 to 0.386 than the unweighted complete case estimate (0.277 (0.255, 0.299)).

## DISCUSSION

We investigated the combined impact of missing data and selection bias on association estimates using simulation studies and real-world EHR data. Our study shows that biobanks with genetic data can reduce these biases by incorporating genetic summaries of exposures and outcomes. Building on previous research,^3,51^ we assessed the effectiveness of PRS-informed multiple imputation in improving association estimates, and to examine the interaction between multiple imputation and sample weighting using simulations and a case study. To our knowledge, this is the first paper to explore the joint impacts of genetic-informed multiple imputation and sample weighting methods in EHR-linked biobank data.

Our simulations revealed that PRS-informed multiple imputation generally outperformed standard methods, particularly for MAR and MNAR data, by offering smaller confidence intervals (Supplementary Figure 3) and reduced bias. PRS preserve correlation between underlying traits despite often being weakly predictive of the trait itself.^51^ Because PRS are observed on a large fraction of the sample, they may help with selection and non-response biases. However, while PRS-imputed analyses improved coverage rates and reduced bias compared to complete case analyses, as expected, ^26,65^ they did not fully recover the nominal coverage rate, especially under MNAR conditions. Notably, PRS-imputed analyses also demonstrated the lowest RMSE in MAR scenarios, suggesting better estimation accuracy.

Missingness in EHR-linked biobank data often deviate from the MCAR assumption due to factors such as patient health status, healthcare access, and EHR fragmentation, leading to biased observation processes.^2,4,6,35,47,72–77^ PRS-informed imputation performed best but struggled to achieve nominal coverage or bias reduction when data were MNAR. While correlations between exposures and their PRS are weak (Supplementary Figure 1), stronger correlates would likely improve multiple imputation.

EHR-linked biobank data are subject to selection bias, including due to healthy volunteer bias (as in the UK Biobank^57^) or non-random recruitment strategies (as in MGI^55^ and the NIH All of Us Research Program^56^). When simulating selection bias by oversampling by age, BMI, and glucose, all methods showed substantial bias in unweighted analyses (≥21%). Weighting improved PRS-imputed analyses’ performance, especially for MAR data, significantly reducing bias and nearly restoring nominal coverage rates (e.g., woPRS-imputed vs. PRS-imputed coverage rate for n=1,000: 0.842 vs. 0.840; n=10,000: 0.884 vs. 0.906). PRS-imputed methods only slightly improved bias and RMSE for MNAR data (e.g., n=1,000: 33.67% vs. 32.40%; n=10,000: 25.46% vs. 23.19%).

In the case study using MGI data, we estimated the BMI coefficient for glucose and found small differences between complete case and imputed estimates, likely due to low levels of missingness (non-Hispanic Whites and Blacks: glucose: 14% and 13%; BMI: 0.5% and 1.1%). However, accounting for selection bias resulted in more substantial changes, underscoring its greater impact than missing data.

Our findings suggest that while PRS-informed multiple imputation can enhance the accuracy of association estimates, particularly in MAR scenarios, it does not fully address challenges when data are MNAR. Sensitivity analyses, alongside expert knowledge^78,79^ and tools like m-graphs or m-DAGs,^80–83^ are recommended and methods like Heckman imputation^84,85^ and pattern-mixture models^86,87^ can be explored. For most regression models, complete case analyses can give unbiased results when the probability of being a complete case is independent of the outcome after taking covariates into account, regardless of the missingness mechanism (Supplementary Table 9).^88–90^ Combining PRS-informed multiple imputation with sampling weights can reduce bias and improve coverage, but careful consideration of underlying data-generating mechanisms is essential.

### Strengths and limitations

This study emphasizes the need to address missing data and selection bias in EHR-linked biobanks and suggests actions for researchers. Our simulations highlight the effectiveness of multiple imputation combined with weighting methods. However, our study has limitations. First, our simulations considered a single level of missingness (∼25%) in two scenarios: exposure alone and exposure and outcome, whereas in practice, multiple patterns and levels of missingness can affect exposures, outcomes, and covariates simultaneously. Future studies should explore a wider range of missingness scenarios. Second, selection bias varies across EHR-linked biobanks due to differing recruitment strategies. For instance, MGI has notable selection biases relative to the US adult population, which may not be as pronounced in population-based biobanks like the NIH All of Us Research Program or the UK Biobank. Third, our case study examined a single association parameter with a relatively small level of missingness and without a gold standard estimate. Future research should investigate associations where gold standard estimates are available. Fourth, the study focused on glucose levels, which can be collected without fasting conditions and managed with medication, complicating interpretation. Codes specifying fasting conditions were rarely used and thus not considered in our analyses, and we did not consider other factors that might impact glucose levels (e.g., surgery, metformin use in people with pre-diabetes). Fifth, we examined the association between two continuous variables after collapsing longitudinal data, whereas many studies utilize binary outcomes and longitudinal data. Future work should address these data types. Lastly, clinically informative visiting processes in EHR data increase the likelihood of MNAR data. Although PRS-imputed analyses showed some improvements for MNAR data, future research should incorporate methods that specifically model these processes.^91–94^

### Conclusion

Missing data is a critical issue in EHR-linked biobank data. We leveraged non-missing genetic data – a key feature of biobanks – to assess if PRS-informed multiple imputation could reduce bias in association estimation. Our simulations demonstrated a substantial reduction in bias for MAR data when incorporating genetic information. Using real-world MGI data, selection bias was relatively more impactful than missing data. Our findings call for exploring additional missingness patterns and levels across associations. Biobanks should provide PRS for common exposures available as proxies and sampling weights to address selection bias. This approach will help researchers better mitigate multiple biases in EHR-linked biobank association analyses, enhancing the reliability and validity of their findings.

## ACKNOWLEDGMENTS

Michigan Genomics Initiative: The authors acknowledge the Michigan Genomics Initiative participants, Precision Health at the University of Michigan, the University of Michigan Medical School Central Biorepository, the University of Michigan Medical School Data Office for Clinical and Translational Research, and the University of Michigan Advanced Genomics Core for providing data and specimen storage, management, processing, and distribution services, and the Center for Statistical Genetics in the Department of Biostatistics at the School of Public Health for genotype data curation, imputation, and management in support of the research reported in this publication/grant application/presentation.

## Data-availability statement

Patient confidentiality prevents the sharing of data publicly. However, the data underlying the study’s results are available from the Michigan Genomics Initiative at https://precisionhealth.umich.edu/ourresearch/michigangenomics/ for researchers who meet the criteria for confidential data access. The code used to conduct analyses in this paper is publicly available at https://github.com/maxsal/exprs_imputation.

## Institutional approval

The institutional review board of the University of Michigan Medical School gave ethical approval for this work (HUM00155849).

