## Supplementary Materials for "Reducing Information and Selection Bias in EHR-Linked Biobanks via Genetics-Informed Multiple Imputation and Sample Weighting"

*Supplementary material to…*

Impact of Polygenic Risk Score-Informed Multiple Imputation and Sample Weighting for Handling Missing Data and Selection Bias on Association Estimation in EHR-Linked Biobanks

Maxwell Salvatore, Ritoban Kundu, Jiacong Du, Christopher R Friese, Alison M Mondul, David Hanauer, Haidong Lu, Celeste Leigh Pearce, Bhramar Mukherjee

Supplementary Figure 1 Pairwise complete observation correlation matrix of relevant variables observed in MGI.

Supplementary Table 1 Intercept values for exposure only and exposure and outcome missingness generation models by missing data mechanism.

|  |  | **Exposure and outcome** | | **Exposure only** |
| --- | --- | --- | --- | --- |
| **Mechanisms** | **n** | Exposure | Outcome | Exposure |
| MAR | 1,000 | -5.53 | -4.09 | -5.58 |
|  | 2,500 | -5.36 | -4.02 | -5.36 |
|  | 5,000 | -5.15 | -3.90 | -5.15 |
|  | 10,000 | -4.88 | -3.79 | -4.88 |
|  | **n** | Exposure | Outcome | Exposure |
| MNAR | 1,000 | -7.36 | -7.36 | -7.41 |
|  | 2,500 | -7.05 | -7.05 | -7.04 |
|  | 5,000 | -6.74 | -6.74 | -6.72 |
|  | 10,000 | -6.32 | -6.32 | -6.31 |

Supplementary Table 2 Performance of missing data methods for estimating covariate-adjusted BMI coefficient for glucose by missing data mechanism, metric, and sample size in random sampling simulations with exposure only missingness.

|  |  |  | **Sample size** | | | |
| --- | --- | --- | --- | --- | --- | --- |
| **Mechanism** | **Metric** | **Method** | **1,000** | **2,500** | **5,000** | **10,000** |
| MCAR | Percent bias | Complete case | 0.613 | 0.668 | 0.054 | 0.157 |
|  |  | woPRS-imputed | 0.773 | 0.806 | 0.095 | 0.254 |
|  |  | PRS-imputed | 0.807 | 0.811 | 0.071 | 0.125 |
|  | Coverage rate | Complete case | 0.956 | 0.961 | 0.947 | 0.956 |
|  |  | woPRS-imputed | 0.958 | 0.958 | 0.945 | 0.949 |
|  |  | PRS-imputed | 0.949 | 0.963 | 0.943 | 0.959 |
|  | Average width | Complete case | 0.135 | 0.085 | 0.060 | 0.042 |
|  |  | woPRS-imputed | 0.139 | 0.088 | 0.062 | 0.044 |
|  |  | PRS-imputed | 0.138 | 0.087 | 0.061 | 0.043 |
|  | RMSE | Complete case | 0.033 | 0.021 | 0.016 | 0.011 |
|  |  | woPRS-imputed | 0.033 | 0.022 | 0.016 | 0.011 |
|  |  | PRS-imputed | 0.033 | 0.022 | 0.016 | 0.011 |
| MAR | Percent bias | Complete case | 8.861 | 8.231 | 8.240 | 7.801 |
|  |  | woPRS-imputed | 2.892 | 0.904 | 0.736 | 0.100 |
|  |  | PRS-imputed | 1.538 | 0.555 | 0.465 | 0.041 |
|  | Coverage rate | Complete case | 0.916 | 0.887 | 0.798 | 0.689 |
|  |  | woPRS-imputed | 0.931 | 0.919 | 0.926 | 0.924 |
|  |  | PRS-imputed | 0.951 | 0.951 | 0.940 | 0.945 |
|  | Average width | Complete case | 0.129 | 0.081 | 0.058 | 0.041 |
|  |  | woPRS-imputed | 0.147 | 0.093 | 0.065 | 0.045 |
|  |  | PRS-imputed | 0.143 | 0.091 | 0.064 | 0.045 |
|  | RMSE | Complete case | 0.038 | 0.026 | 0.022 | 0.019 |
|  |  | woPRS-imputed | 0.040 | 0.025 | 0.017 | 0.012 |
|  |  | PRS-imputed | 0.036 | 0.022 | 0.016 | 0.011 |
| MNAR | Percent bias | Complete case | 41.080 | 41.174 | 40.905 | 40.615 |
|  |  | woPRS-imputed | 36.115 | 35.437 | 34.986 | 34.598 |
|  |  | PRS-imputed | 31.946 | 31.670 | 31.391 | 31.074 |
|  | Coverage rate | Complete case | 0.337 | 0.035 | 0.002 | 0.000 |
|  |  | woPRS-imputed | 0.561 | 0.217 | 0.034 | 0.000 |
|  |  | PRS-imputed | 0.637 | 0.278 | 0.055 | 0.001 |
|  | Average width | Complete case | 0.136 | 0.086 | 0.061 | 0.043 |
|  |  | woPRS-imputed | 0.159 | 0.101 | 0.071 | 0.050 |
|  |  | PRS-imputed | 0.156 | 0.098 | 0.069 | 0.049 |
|  | RMSE | Complete case | 0.089 | 0.084 | 0.083 | 0.081 |
|  |  | woPRS-imputed | 0.082 | 0.074 | 0.072 | 0.070 |
|  |  | PRS-imputed | 0.074 | 0.067 | 0.065 | 0.063 |
| Abbreviations: BMI, body mass index; CC, complete case; MAR, missing at random; MCAR, missing completely at random; MNAR, missing not at random; PRS, polygenic risk score; RMSE, root mean square error; woPRS, without polygenic risk score | | | | | | |

Supplementary Table 3 Performance of missing data methods for estimating covariate-adjusted BMI coefficient for glucose by missing data mechanism, metric, and sample size in random sampling simulations with exposure and outcome missingness.

|  |  |  | **Sample size** | | | |
| --- | --- | --- | --- | --- | --- | --- |
| **Mechanism** | **Metric** | **Method** | **1,000** | **2,500** | **5,000** | **10,000** |
| MCAR | Percent bias | Complete case | 0.320 | 0.234 | 0.125 | 0.217 |
|  |  | woPRS-imputed | 0.628 | 0.256 | 0.286 | 0.151 |
|  |  | PRS-imputed | 0.109 | 0.429 | 0.120 | 0.043 |
|  | Coverage rate | Complete case | 0.938 | 0.956 | 0.961 | 0.951 |
|  |  | woPRS-imputed | 0.939 | 0.950 | 0.946 | 0.956 |
|  |  | PRS-imputed | 0.941 | 0.951 | 0.956 | 0.951 |
|  | Average width | Complete case | 0.156 | 0.098 | 0.069 | 0.049 |
|  |  | woPRS-imputed | 0.170 | 0.107 | 0.076 | 0.054 |
|  |  | PRS-imputed | 0.167 | 0.107 | 0.074 | 0.053 |
|  | RMSE | Complete case | 0.042 | 0.025 | 0.017 | 0.012 |
|  |  | woPRS-imputed | 0.043 | 0.026 | 0.018 | 0.013 |
|  |  | PRS-imputed | 0.042 | 0.026 | 0.018 | 0.012 |
| MAR | Percent bias | Complete case | 7.021 | 7.506 | 7.489 | 7.009 |
|  |  | woPRS-imputed | 2.061 | 2.140 | 1.828 | 1.319 |
|  |  | PRS-imputed | 1.877 | 1.395 | 1.364 | 0.968 |
|  | Coverage rate | Complete case | 0.925 | 0.914 | 0.857 | 0.784 |
|  |  | woPRS-imputed | 0.929 | 0.952 | 0.945 | 0.946 |
|  |  | PRS-imputed | 0.930 | 0.946 | 0.948 | 0.947 |
|  | Average width | Complete case | 0.145 | 0.091 | 0.064 | 0.046 |
|  |  | woPRS-imputed | 0.180 | 0.113 | 0.078 | 0.056 |
|  |  | PRS-imputed | 0.166 | 0.104 | 0.075 | 0.053 |
|  | RMSE | Complete case | 0.041 | 0.028 | 0.022 | 0.018 |
|  |  | woPRS-imputed | 0.044 | 0.027 | 0.019 | 0.013 |
|  |  | PRS-imputed | 0.042 | 0.026 | 0.018 | 0.013 |
| MNAR | Percent bias | Complete case | 62.273 | 62.237 | 62.420 | 62.177 |
|  |  | woPRS-imputed | 55.679 | 54.781 | 54.450 | 54.180 |
|  |  | PRS-imputed | 53.889 | 53.148 | 53.282 | 52.867 |
|  | Coverage rate | Complete case | 0.117 | 0.000 | 0.000 | 0.000 |
|  |  | woPRS-imputed | 0.296 | 0.025 | 0.001 | 0.000 |
|  |  | PRS-imputed | 0.299 | 0.022 | 0.002 | 0.000 |
|  | Average width | Complete case | 0.149 | 0.094 | 0.066 | 0.047 |
|  |  | woPRS-imputed | 0.172 | 0.109 | 0.076 | 0.053 |
|  |  | PRS-imputed | 0.167 | 0.107 | 0.074 | 0.052 |
|  | RMSE | Complete case | 0.130 | 0.126 | 0.125 | 0.124 |
|  |  | woPRS-imputed | 0.118 | 0.112 | 0.109 | 0.108 |
|  |  | PRS-imputed | 0.115 | 0.108 | 0.107 | 0.106 |
| Abbreviations: BMI, body mass index; CC, complete case; MAR, missing at random; MCAR, missing completely at random; MNAR, missing not at random; PRS, polygenic risk score; RMSE, root mean square error; woPRS, without polygenic risk score | | | | | | |

Supplementary Table 4 Biased/covariate-informed sampling simulation performance of missing data methods for estimating unweighted and covariate-adjusted BMI coefficient for glucose by missing data mechanism, metric, and sample size with exposure only missingness.

|  |  |  | **Sample size** | | | |
| --- | --- | --- | --- | --- | --- | --- |
| **Mechanism** | **Metric** | **Method** | **1,000** | **2,500** | **5,000** | **10,000** |
| MCAR | Percent bias | Complete case | 36.873 | 51.226 | 63.133 | 73.374 |
|  |  | woPRS-imputed | 29.057 | 44.652 | 58.862 | 71.132 |
|  |  | PRS-imputed | 35.796 | 50.593 | 62.758 | 73.435 |
|  | Coverage rate | Complete case | 0.456 | 0.005 | 0.000 | 0.000 |
|  |  | woPRS-imputed | 0.690 | 0.065 | 0.001 | 0.000 |
|  |  | PRS-imputed | 0.500 | 0.013 | 0.000 | 0.000 |
|  | Average width | Complete case | 0.137 | 0.087 | 0.062 | 0.043 |
|  |  | woPRS-imputed | 0.155 | 0.099 | 0.071 | 0.050 |
|  |  | PRS-imputed | 0.142 | 0.090 | 0.064 | 0.046 |
|  | RMSE | Complete case | 0.081 | 0.104 | 0.126 | 0.146 |
|  |  | woPRS-imputed | 0.070 | 0.092 | 0.118 | 0.142 |
|  |  | PRS-imputed | 0.080 | 0.103 | 0.125 | 0.146 |
| MAR | Percent bias | Complete case | 34.998 | 48.445 | 60.272 | 71.844 |
|  |  | woPRS-imputed | 39.082 | 54.004 | 69.995 | 85.814 |
|  |  | PRS-imputed | 41.165 | 55.207 | 68.433 | 81.637 |
|  | Coverage rate | Complete case | 0.452 | 0.005 | 0.000 | 0.000 |
|  |  | woPRS-imputed | 0.659 | 0.103 | 0.000 | 0.000 |
|  |  | PRS-imputed | 0.450 | 0.014 | 0.001 | 0.000 |
|  | Average width | Complete case | 0.132 | 0.083 | 0.059 | 0.042 |
|  |  | woPRS-imputed | 0.186 | 0.121 | 0.083 | 0.058 |
|  |  | PRS-imputed | 0.154 | 0.097 | 0.070 | 0.050 |
|  | RMSE | Complete case | 0.077 | 0.098 | 0.120 | 0.143 |
|  |  | woPRS-imputed | 0.092 | 0.113 | 0.142 | 0.171 |
|  |  | PRS-imputed | 0.091 | 0.113 | 0.137 | 0.163 |
| MNAR | Percent bias | Complete case | 67.098 | 80.842 | 93.566 | 106.255 |
|  |  | woPRS-imputed | 84.533 | 99.765 | 113.517 | 127.188 |
|  |  | PRS-imputed | 77.451 | 91.727 | 105.719 | 119.905 |
|  | Coverage rate | Complete case | 0.034 | 0.000 | 0.000 | 0.000 |
|  |  | woPRS-imputed | 0.085 | 0.000 | 0.000 | 0.000 |
|  |  | PRS-imputed | 0.052 | 0.000 | 0.000 | 0.000 |
|  | Average width | Complete case | 0.138 | 0.087 | 0.062 | 0.044 |
|  |  | woPRS-imputed | 0.177 | 0.109 | 0.075 | 0.052 |
|  |  | PRS-imputed | 0.160 | 0.101 | 0.072 | 0.050 |
|  | RMSE | Complete case | 0.138 | 0.162 | 0.186 | 0.211 |
|  |  | woPRS-imputed | 0.175 | 0.200 | 0.226 | 0.253 |
|  |  | PRS-imputed | 0.160 | 0.184 | 0.211 | 0.238 |
| Abbreviations: BMI, body mass index; CC, complete case; MAR, missing at random; MCAR, missing completely at random; MNAR, missing not at random; PRS, polygenic risk score; RMSE, root mean square error; woPRS, without polygenic risk score | | | | | | |

Supplementary Table 5 Biased/covariate-informed sampling simulation performance of missing data methods for estimating weighted and covariate-adjusted BMI coefficient for glucose by missing data mechanism, metric, and sample size with exposure only missingness.

|  |  |  | **Sample size** | | | |
| --- | --- | --- | --- | --- | --- | --- |
| **Mechanism** | **Metric** | **Method** | **1,000** | **2,500** | **5,000** | **10,000** |
| MCAR | Percent bias | Complete case | 32.506 | 17.254 | 12.133 | 6.518 |
|  |  | woPRS-imputed | 1.842 | 11.046 | 11.583 | 12.559 |
|  |  | PRS-imputed | 6.144 | 3.518 | 5.327 | 7.055 |
|  | Coverage rate | Complete case | 0.709 | 0.773 | 0.792 | 0.821 |
|  |  | woPRS-imputed | 0.874 | 0.899 | 0.926 | 0.925 |
|  |  | PRS-imputed | 0.859 | 0.907 | 0.931 | 0.941 |
|  | Average width | Complete case | 0.356 | 0.286 | 0.229 | 0.177 |
|  |  | woPRS-imputed | 0.346 | 0.266 | 0.208 | 0.163 |
|  |  | PRS-imputed | 0.349 | 0.270 | 0.213 | 0.165 |
|  | RMSE | Complete case | 0.157 | 0.108 | 0.084 | 0.061 |
|  |  | woPRS-imputed | 0.112 | 0.084 | 0.065 | 0.052 |
|  |  | PRS-imputed | 0.114 | 0.081 | 0.063 | 0.048 |
| MAR | Percent bias | Complete case | 32.936 | 19.958 | 15.178 | 10.345 |
|  |  | woPRS-imputed | 11.881 | 0.514 | 2.535 | 4.511 |
|  |  | PRS-imputed | 14.274 | 2.992 | 0.715 | 3.800 |
|  | Coverage rate | Complete case | 0.693 | 0.747 | 0.766 | 0.784 |
|  |  | woPRS-imputed | 0.779 | 0.831 | 0.866 | 0.877 |
|  |  | PRS-imputed | 0.774 | 0.828 | 0.867 | 0.883 |
|  | Average width | Complete case | 0.343 | 0.274 | 0.219 | 0.173 |
|  |  | woPRS-imputed | 0.327 | 0.258 | 0.205 | 0.160 |
|  |  | PRS-imputed | 0.328 | 0.259 | 0.206 | 0.161 |
|  | RMSE | Complete case | 0.151 | 0.107 | 0.082 | 0.063 |
|  |  | woPRS-imputed | 0.129 | 0.093 | 0.071 | 0.056 |
|  |  | PRS-imputed | 0.130 | 0.093 | 0.070 | 0.055 |
| MNAR | Percent bias | Complete case | 40.836 | 28.779 | 25.531 | 23.222 |
|  |  | woPRS-imputed | 29.135 | 17.697 | 14.954 | 14.002 |
|  |  | PRS-imputed | 29.067 | 17.204 | 14.151 | 12.657 |
|  | Coverage rate | Complete case | 0.662 | 0.705 | 0.702 | 0.639 |
|  |  | woPRS-imputed | 0.705 | 0.754 | 0.766 | 0.739 |
|  |  | PRS-imputed | 0.705 | 0.758 | 0.770 | 0.761 |
|  | Average width | Complete case | 0.347 | 0.278 | 0.224 | 0.179 |
|  |  | woPRS-imputed | 0.337 | 0.269 | 0.215 | 0.170 |
|  |  | PRS-imputed | 0.336 | 0.268 | 0.214 | 0.170 |
|  | RMSE | Complete case | 0.160 | 0.117 | 0.093 | 0.077 |
|  |  | woPRS-imputed | 0.145 | 0.103 | 0.080 | 0.065 |
|  |  | PRS-imputed | 0.145 | 0.103 | 0.079 | 0.063 |
| Abbreviations: BMI, body mass index; CC, complete case; MAR, missing at random; MCAR, missing completely at random; MNAR, missing not at random; PRS, polygenic risk score; RMSE, root mean square error; woPRS, without polygenic risk score | | | | | | |

Supplementary Table 6 Biased/covariate-informed sampling simulation performance of missing data methods for estimating unweighted and covariate-adjusted BMI coefficient for glucose by missing data mechanism, metric, and sample size with exposure and outcome missingness.

|  |  |  | **Sample size** | | | |
| --- | --- | --- | --- | --- | --- | --- |
| **Mechanism** | **Metric** | **Method** | **1,000** | **2,500** | **5,000** | **10,000** |
| MCAR | Percent bias | Complete case | 37.159 | 51.197 | 63.088 | 73.468 |
|  |  | woPRS-imputed | 21.696 | 34.714 | 49.350 | 64.485 |
|  |  | PRS-imputed | 31.311 | 44.849 | 57.490 | 69.527 |
|  | Coverage rate | Complete case | 0.563 | 0.027 | 0.000 | 0.000 |
|  |  | woPRS-imputed | 0.850 | 0.476 | 0.077 | 0.022 |
|  |  | PRS-imputed | 0.703 | 0.147 | 0.003 | 0.000 |
|  | Average width | Complete case | 0.159 | 0.100 | 0.071 | 0.050 |
|  |  | woPRS-imputed | 0.196 | 0.132 | 0.107 | 0.092 |
|  |  | PRS-imputed | 0.180 | 0.114 | 0.084 | 0.061 |
|  | RMSE | Complete case | 0.084 | 0.105 | 0.126 | 0.146 |
|  |  | woPRS-imputed | 0.068 | 0.077 | 0.103 | 0.132 |
|  |  | PRS-imputed | 0.078 | 0.094 | 0.116 | 0.139 |
| MAR | Percent bias | Complete case | 32.508 | 46.041 | 58.330 | 70.157 |
|  |  | woPRS-imputed | 38.110 | 50.608 | 69.346 | 90.519 |
|  |  | PRS-imputed | 40.978 | 53.365 | 68.150 | 83.145 |
|  | Coverage rate | Complete case | 0.596 | 0.026 | 0.000 | 0.000 |
|  |  | woPRS-imputed | 0.754 | 0.306 | 0.025 | 0.003 |
|  |  | PRS-imputed | 0.579 | 0.080 | 0.000 | 0.000 |
|  | Average width | Complete case | 0.148 | 0.093 | 0.066 | 0.047 |
|  |  | woPRS-imputed | 0.223 | 0.149 | 0.114 | 0.083 |
|  |  | PRS-imputed | 0.185 | 0.117 | 0.087 | 0.062 |
|  | RMSE | Complete case | 0.075 | 0.094 | 0.117 | 0.140 |
|  |  | woPRS-imputed | 0.102 | 0.111 | 0.145 | 0.184 |
|  |  | PRS-imputed | 0.098 | 0.111 | 0.138 | 0.167 |
| MNAR | Percent bias | Complete case | 85.115 | 98.036 | 110.564 | 124.209 |
|  |  | woPRS-imputed | 120.797 | 135.571 | 149.429 | 164.005 |
|  |  | PRS-imputed | 109.921 | 122.884 | 138.468 | 155.203 |
|  | Coverage rate | Complete case | 0.014 | 0.000 | 0.000 | 0.000 |
|  |  | woPRS-imputed | 0.064 | 0.002 | 0.000 | 0.000 |
|  |  | PRS-imputed | 0.032 | 0.000 | 0.000 | 0.000 |
|  | Average width | Complete case | 0.152 | 0.096 | 0.068 | 0.048 |
|  |  | woPRS-imputed | 0.217 | 0.141 | 0.094 | 0.063 |
|  |  | PRS-imputed | 0.195 | 0.122 | 0.086 | 0.060 |
|  | RMSE | Complete case | 0.173 | 0.196 | 0.220 | 0.247 |
|  |  | woPRS-imputed | 0.249 | 0.272 | 0.298 | 0.326 |
|  |  | PRS-imputed | 0.227 | 0.247 | 0.277 | 0.309 |
| Abbreviations: BMI, body mass index; CC, complete case; MAR, missing at random; MCAR, missing completely at random; MNAR, missing not at random; PRS, polygenic risk score; RMSE, root mean square error; woPRS, without polygenic risk score | | | | | | |

Supplementary Table 7 Biased/covariate-informed sampling simulation performance of missing data methods for estimating weighted and covariate-adjusted BMI coefficient for glucose by missing data mechanism, metric, and sample size with exposure and outcome missingness.

|  |  |  | **Sample size** | | | |
| --- | --- | --- | --- | --- | --- | --- |
| **Mechanism** | **Metric** | **Method** | **1,000** | **2,500** | **5,000** | **10,000** |
| MCAR | Percent bias | Complete case | 37.174 | 20.417 | 14.551 | 8.517 |
|  |  | woPRS-imputed | 25.938 | 33.332 | 31.954 | 28.911 |
|  |  | PRS-imputed | 17.280 | 25.151 | 24.982 | 24.115 |
|  | Coverage rate | Complete case | 0.684 | 0.761 | 0.791 | 0.810 |
|  |  | woPRS-imputed | 0.863 | 0.831 | 0.793 | 0.748 |
|  |  | PRS-imputed | 0.890 | 0.896 | 0.875 | 0.834 |
|  | Average width | Complete case | 0.375 | 0.303 | 0.246 | 0.189 |
|  |  | woPRS-imputed | 0.381 | 0.281 | 0.222 | 0.169 |
|  |  | PRS-imputed | 0.384 | 0.289 | 0.228 | 0.171 |
|  | RMSE | Complete case | 0.172 | 0.119 | 0.090 | 0.066 |
|  |  | woPRS-imputed | 0.123 | 0.104 | 0.087 | 0.073 |
|  |  | PRS-imputed | 0.115 | 0.093 | 0.078 | 0.065 |
| MAR | Percent bias | Complete case | 35.885 | 21.217 | 16.427 | 10.861 |
|  |  | woPRS-imputed | 1.039 | 8.231 | 9.369 | 7.404 |
|  |  | PRS-imputed | 3.671 | 5.991 | 7.541 | 7.380 |
|  | Coverage rate | Complete case | 0.682 | 0.754 | 0.769 | 0.776 |
|  |  | woPRS-imputed | 0.842 | 0.863 | 0.890 | 0.884 |
|  |  | PRS-imputed | 0.840 | 0.860 | 0.895 | 0.906 |
|  | Average width | Complete case | 0.360 | 0.289 | 0.232 | 0.185 |
|  |  | woPRS-imputed | 0.350 | 0.271 | 0.213 | 0.166 |
|  |  | PRS-imputed | 0.348 | 0.273 | 0.214 | 0.166 |
|  | RMSE | Complete case | 0.160 | 0.114 | 0.087 | 0.068 |
|  |  | woPRS-imputed | 0.125 | 0.092 | 0.072 | 0.058 |
|  |  | PRS-imputed | 0.124 | 0.092 | 0.069 | 0.055 |
| MNAR | Percent bias | Complete case | 49.811 | 37.496 | 34.837 | 33.700 |
|  |  | woPRS-imputed | 33.674 | 23.788 | 23.089 | 25.463 |
|  |  | PRS-imputed | 32.404 | 21.942 | 21.067 | 23.194 |
|  | Coverage rate | Complete case | 0.627 | 0.655 | 0.636 | 0.542 |
|  |  | woPRS-imputed | 0.685 | 0.723 | 0.705 | 0.599 |
|  |  | PRS-imputed | 0.695 | 0.725 | 0.720 | 0.621 |
|  | Average width | Complete case | 0.357 | 0.287 | 0.233 | 0.189 |
|  |  | woPRS-imputed | 0.339 | 0.270 | 0.217 | 0.172 |
|  |  | PRS-imputed | 0.338 | 0.269 | 0.216 | 0.171 |
|  | RMSE | Complete case | 0.174 | 0.129 | 0.108 | 0.094 |
|  |  | woPRS-imputed | 0.151 | 0.110 | 0.089 | 0.079 |
|  |  | PRS-imputed | 0.149 | 0.108 | 0.087 | 0.076 |
| Abbreviations: BMI, body mass index; CC, complete case; MAR, missing at random; MCAR, missing completely at random; MNAR, missing not at random; PRS, polygenic risk score; RMSE, root mean square error; woPRS, without polygenic risk score | | | | | | |

Supplementary Figure 2 Average 95% CI width (panels A and B) and RMSE (panels C and D) diagnostics for BMI coefficient for glucose by missing data mechanism and method and sample size under random sampling with exposure only (panels A and C) and exposure and outcome missingness (panels B and D).

Analyses were adjusted for age, sex, non-Hispanic White, and smoking status (ever/never). Corresponding coverage rate, percent bias, average confidence interval width, and root mean squared error diagnostics are reported in Supplementary Table 2 and Supplementary Table 3 . Abbreviations: MAR, missing at random; MCAR, missing completely at random; MNAR, missing not at random; PRS, polygenic risk score; RMSE, root mean square error; woPRS, without polygenic risk score.

Supplementary Figure 3 Average 95% CI width for unweighted (left) and weighted (right) BMI coefficient for glucose by missing data mechanism and method and sample size under biased sampling with exposure only (panel A) and exposure and outcome missingness (panel B).

For biased sampling simulations, unweighted and weighted diagnostics are reported in Supplementary Table 4 , Supplementary Table 5 , Supplementary Table 6 , and Supplementary Table 7 . Analyses were adjusted for age, sex, non-Hispanic White, and smoking status (ever/never). Abbreviations: MAR, missing at random; MCAR, missing completely at random; MNAR, missing not at random; PRS, polygenic risk score; RMSE, root mean square error; woPRS, without polygenic risk score.

Supplementary Figure 4 RMSE for unweighted (left) and weighted (right) BMI coefficient for glucose by missing data mechanism and method and sample size under biased sampling with exposure only (panel A) and exposure and outcome missingness (panel B).

For biased sampling simulations, unweighted and weighted diagnostics are reported in Supplementary Table 4 , Supplementary Table 5 , Supplementary Table 6 , and Supplementary Table 7 . Analyses were adjusted for age, sex, non-Hispanic White, and smoking status (ever/never). Abbreviations: MAR, missing at random; MCAR, missing completely at random; MNAR, missing not at random; PRS, polygenic risk score; woPRS, without polygenic risk score.

Supplementary Table 8 Comparison of demographic, health measurements, and polygenic risk score values overall and among adults 40 or older without diabetes, with and without any missing values in the Michigan Genomics Initiative.

|  | **Overall** | **Incomplete observations** | **Complete observations** |  | **Non-missing PRS** |
| --- | --- | --- | --- | --- | --- |
| **Characteristic** | N = 50,026*^a^* | N = 20,799*^a^* | N = 29,227*^a^* | **p-value*^b^*** | N = 35,353*^a^* |
| Age | 62.9 (12.5) | 61.9 (12.4) | 63.5 (12.5) | <0.001 | 63.1 (12.6) |
| Female | 54.5 (27,261) | 55.1 (11,465) | 54.0 (15,796) | 0.017 | 54.0 (19,100) |
| Non-Hispanic White | 86.0 (42,999) | 84.0 (17,479) | 87.3 (25,520) | <0.001 | 87.5 (30,942) |
| Smoking status (ever) | 48.1 (20,561) | 44.8 (6,071) | 49.6 (14,490) | <0.001 | 49.1 (16,022) |
| *Missing* | *7,238* | *7,238* | *0* |  | *2691* |
| BMI | 29.1 (6.0) | 29.1 (6.0) | 29.0 (6.0) | 0.7 | 29.0 (6.0) |
| *Missing* | *292* | *292* | *0* |  | *156* |
| Glucose | 99.0 (14.1) | 98.8 (14.6) | 99.1 (13.9) | <0.001 | 99.3 (14.2) |
| *Missing* | *5,467* | *5,467* | *0* |  | *3991* |
| BMI PRS^c^ | 0.000 (1.000) | 0.008 (1.023) | -0.002 (0.995) | 0.3 | 0.000 (1.000) |
| *Missing* | *14,673* | *14,673* | *0* |  | *0* |
| Glucose PRS^c^ | 0.000 (1.000) | 0.013 (0.996) | -0.003 (1.001) | 0.2 | 0.000 (1.000) |
| *Missing* | *14,673* | *14,673* | *0* |  | *0* |

Supplementary Figure 5 Estimation of the coefficient for BMI with glucose as the outcome by missing data method and weighting approach among all MGI adults age 40 or older without diabetes (i.e., not stratified by race/ethnicity; n=50,026).

The PRS-imputed subset sample (n=35,353) was restricted to individuals with non-missing genotype data before multiple imputation. Analyses were adjusted for age, sex, smoking status (ever/never), and a non-Hispanic White indicator. Gray shaded regions represent corresponding 95% confidence interval from National Health Interview Survey-weighted All of Us data to make All of Us data more representative of the US population (target population). Abbreviations: PRS, polygenic risk score.

Supplementary Table 9 Potential bias in intercept, exposure, and confounder regression coefficients in complete case analysis of linear and logistic regression by reason for missing data.

|  | Linear regression coefficient | | |  | Logistic regression coefficient | | |
| --- | --- | --- | --- | --- | --- | --- | --- |
| Variables missingness is dependent upon | Intercept | Exposure | Confounder |  | Intercept | Exposure | Confounder |
| None (e.g., missing completely at random) | Unbiased | Unbiased | Unbiased |  | Unbiased | Unbiased | Unbiased |
| Outcome (Y) only | Biased | Biased*^a^* | Biased*^a^* |  | Biased | Unbiased | Unbiased |
| Exposure (X) and/or other covariates (C) | Unbiased | Unbiased | Unbiased |  | Unbiased | Unbiased | Unbiased |
| Outcome (Y) and confounders (C) | Biased | Biased | Biased |  | Biased | Unbiased | Biased |
| Outcome (Y), exposure (X), and possible confounders (C) | Biased | Biased | Biased |  | Biased | Biased*^b^* | Biased |
| *^a^* Biased in general, except when in truth there is no association between the outcome and the exposure or confounding in question (i.e., the true value of the regression coefficient is zero) | | | | | | | |
| *^b^* Biased in general, except when missingness depends on the outcome and exposure independently | | | | | | | |
| Adapted from Supplementary Table 1 from Hughes and colleagues (2019, doi: [10.1093/ije/dyz032](https://www.doi.org/10.1093/ije/dyz032)) and Table 1 from Bartlett and colleagues (2015; doi: [10.1093/aje/kwv114](https://www.doi.org/10.1093/aje/kwv114)) | | | | | | | |
